# Fully automatic summarization of radiology reports using natural language processing with language models

**DOI:** 10.1101/2023.12.01.23299267

**Authors:** Mizuho Nishio, Takaaki Matsunaga, Hidetoshi Matsuo, Munenobu Nogami, Yasuhisa Kurata, Koji Fujimoto, Osamu Sugiyama, Toshiaki Akashi, Shigeki Aoki, Takamichi Murakami

## Abstract

Natural language processing using language models has yielded promising results in various fields. The use of language models may help improve the workflow of radiologists. This retrospective study aimed to construct and evaluate language models for the automatic summarization of radiology reports. Two datasets of radiology reports were used: MIMIC-CXR and the Japan Medical Image Database (JMID). MIMIC-CXR is an open dataset comprising chest radiograph reports. JMID is a large dataset of CT and MRI reports comprising reports from 10 academic medical centers in Japan. A total of 128,032 and 1,101,271 reports from the MIMIC-CXR and JMID, respectively, were included in this study. Four Text-to-Text Transfer Transformer (T5) models were constructed. Recall-Oriented Understudy for Gisting Evaluation (ROUGE), a quantitative metric, was used to evaluate the quality of text summarized from 19,205 and 58,043 test sets from MIMIC-CXR and JMID, respectively. The Wilcoxon signed-rank test was utilized to evaluate the differences among the ROUGE values of the four T5 models. In addition, subsets of automatically summarized text in the test sets were manually evaluated by two radiologists. Based on the Wilcoxon signed-rank test, the best T5 models were selected for the automatic summarization. The quantitative metrics of the best T5 models were as follows: ROUGE-1 = 57.75 ± 30.99, ROUGE-2 = 49.96 ± 35.36, and ROUGE-L = 54.07 ± 32.48 in MIMIC-CXR; ROUGE-1 = 50.00 ± 29.24, ROUGE-2 = 39.66 ± 30.21, and ROUGE-L = 47.87 ± 29.44 in JMID. The radiologists’ evaluations revealed that 86% (86/100) and 85% (85/100) of the texts automatically summarized from MIMIC-CXR and JMID, respectively, were clinically useful. The T5 models constructed in this study were capable of automatic summarization of radiology reports. The radiologists’ evaluations revealed that most of the automatically summarized texts were clinically valuable.

## Introduction

Radiology reports are a valuable source of information for improving clinical practice and supporting research. A multitude of radiology reports have been written in recent years owing to the advances in the field of radiology. However, manually processing a large number of unstructured reports is difficult as radiology reports are often recorded as unstructured data.

Natural language processing (NLP) has enabled computers to process natural languages (1,2) by facilitating the extraction of structured information from electronic medical records and radiology reports. Consequently, NLP has been used for text classification, text summarization, and text generation in the field of radiology (3–5). Recent advances in NLP have been accompanied by the application of deep learning.

NLP has the potential to reduce the workload of radiologists by extracting structured information from radiology reports. This would aid clinicians and radiologists in the decision-making process and in identifying patients for research. However, unlike computer vision (6), NLP has not received significant attention in the field of artificial intelligence, and reviews on the application of NLP in radiology have been limited (5).

The development of language models has been a promising advancement in NLP. Language models are neural networks trained using a large amount of text, and the number of parameters in the model can be used as a measure of performance. Several types of language models have been developed in recent years, such as Bidirectional Encoder Representations from Transformers (7), Text-to-Text Transfer Transformer (T5) (8,9), and Generative Pre-Training-1 (10), Generative Pre-Training-2 (11), Generative Pre-Training-3 (12) and so on (13). These models have achieved state-of-the-art performance in NLP tasks.

Radiology reports are divided into two sections: findings and impression. Automatic summarization of the impression section from the findings section would reduce the workload of radiologists. Thus, this study aimed to investigate the effectiveness of using a language model to summarize radiology reports automatically. The contributions of this study are threefold: (i) The T5 language model was used to summarize radiology reports automatically. (ii) Automatic summarization of radiology reports was performed in two languages: chest radiograph (CXR) reports in English and CT and MRI reports in Japanese. (iii) Recall-Oriented Understudy for Gisting Evaluation (ROUGE), a quantitative evaluation metric, and a semi-quantitative evaluation performed by radiologists were used to evaluate the automatically summarized sentences, and the relationship between the ROUGE metrics and radiologists’ evaluations was investigated.

## Material and method

This retrospective study was approved by the Institutional Review Boards of the Japan Medical Image Database (JMID) project and Kobe University Hospital. The requirement for informed consent was waived. This study was conducted in accordance with the Checklist for Artificial Intelligence in Medical Imaging (14).

### Dataset

Two datasets were used in the study: MIMIC-CXR and JMID. The MIMIC-CXR dataset comprises chest radiographs and the corresponding reports (15), whereas the JMID dataset was obtained from the JMID project, a project wherein 10 academic medical centers in Japan collaborated to create a large radiology database with de-identified patient data. All reports dated from 8/4/2010 to 3/31/2023 were collected from the JMID database. Reports with missing data in the findings or impression sections were excluded. Pairs of findings and impression sections were collected from each report in the two datasets. Figure 1 presents the flow diagrams of the study.

**Figure 1.**
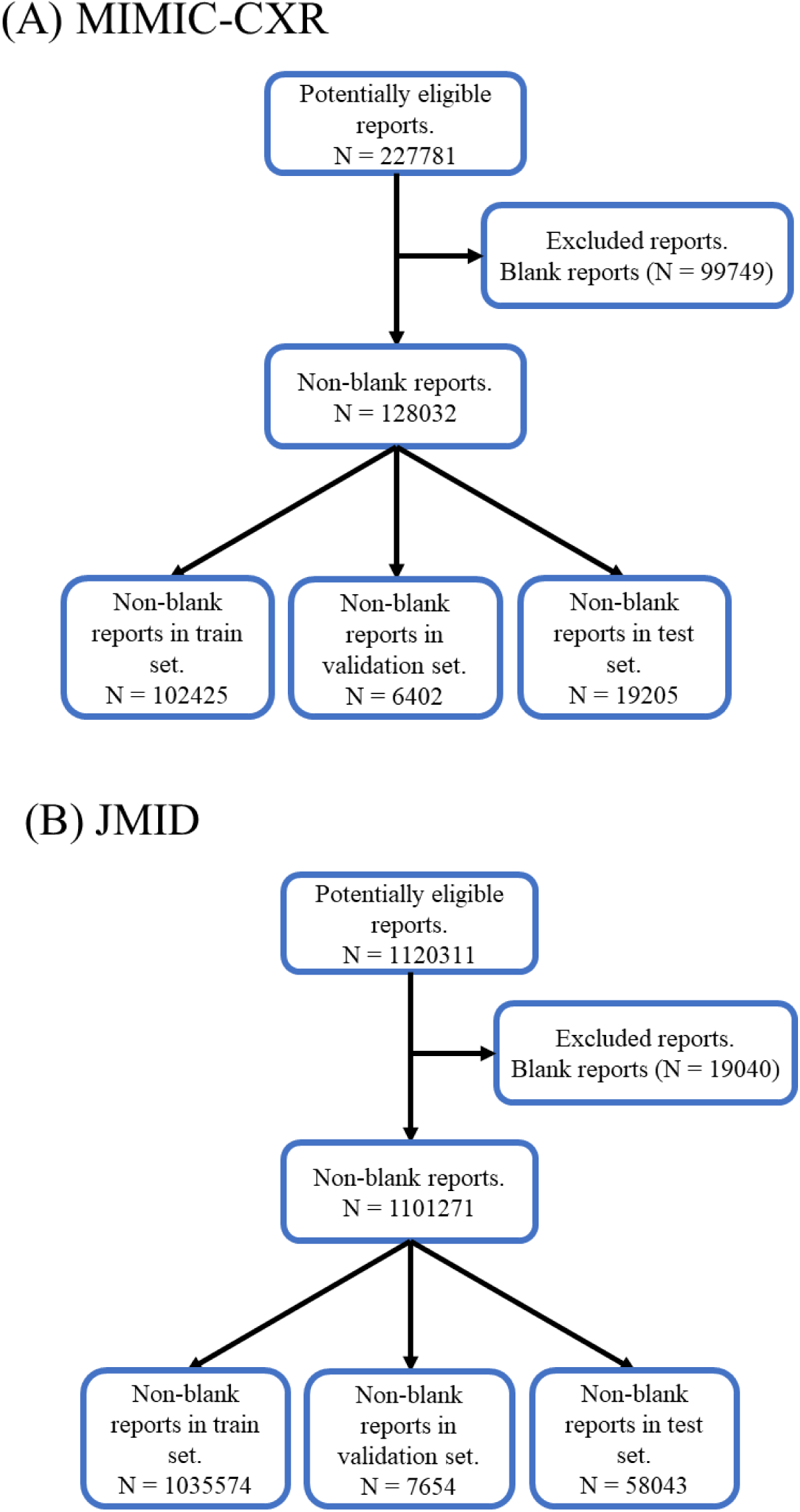
Flowcharts of including radiology reports. (A) MIMIC-CXR, (B) JMID. Abbreviations: JMID, Japan Medical Image Database

### Ground truth

MIMIC-CXR and JMID include clinical radiology reports; thus, the impression sections of actual reports were used as the ground truth.

### Dataset partition

The MIMIC-CXR images were randomly divided into training, validation, and test sets at a 16:1:3 ratio. The reports acquired from JMID were divided into three sets according to the date of the reports: training set, 8/4/2010–11/30/2022 and 12/10/2022–12/31/2022; validation set, 12/1/2022–12/9/2022; and test set, 1/1/2023–3/31/2023. Date-based dataset partitioning was not possible in MIMIC-CXR as the MIMIC-CXR reports were not dated.

### Language model

T5 is a transformer-based neural network model comprising an encoder and decoder (8,9). Fine-tuning the pre-trained T5 model can improve the performance of text summarization, question answering, and text classification (16,17). The present study focused on text summarization (summarization of radiology reports). Consequently, the input was the text of the findings section, whereas the output was that of the impressions section. The T5 model was trained to summarize the findings section automatically via fine-tuning. Pre-trained T5 models were obtained from Hugging Face (https://huggingface.co/models) for fine-tuning. Two pre-trained T5 models (“t5-base” (18) and “google/mt5-base” (19)) were obtained for MIMIC-CXR. Two pre-trained T5 models (“megagonlabs/t5-base-japanese-web” (20) and “google/mt5-base” (19)) were obtained for JMID. “t5-base,” “megagonlabs/t5-base-japanese-web,” and “google/mt5-base” are the pre-trained English, Japanese, and multilingual models, respectively.

### Model training

Figure 2 summarizes the process of model development and prediction using the T5 model. Fine-tuning of the T5 models is detailed in the Appendix 1. Batch sizes of 2 and 8 were used for fine-tuning. There were two choices of pre-trained models and two types of batch sizes for MIMIC-CXR and JMID, resulting in a total of four combinations.

**Figure 2.**
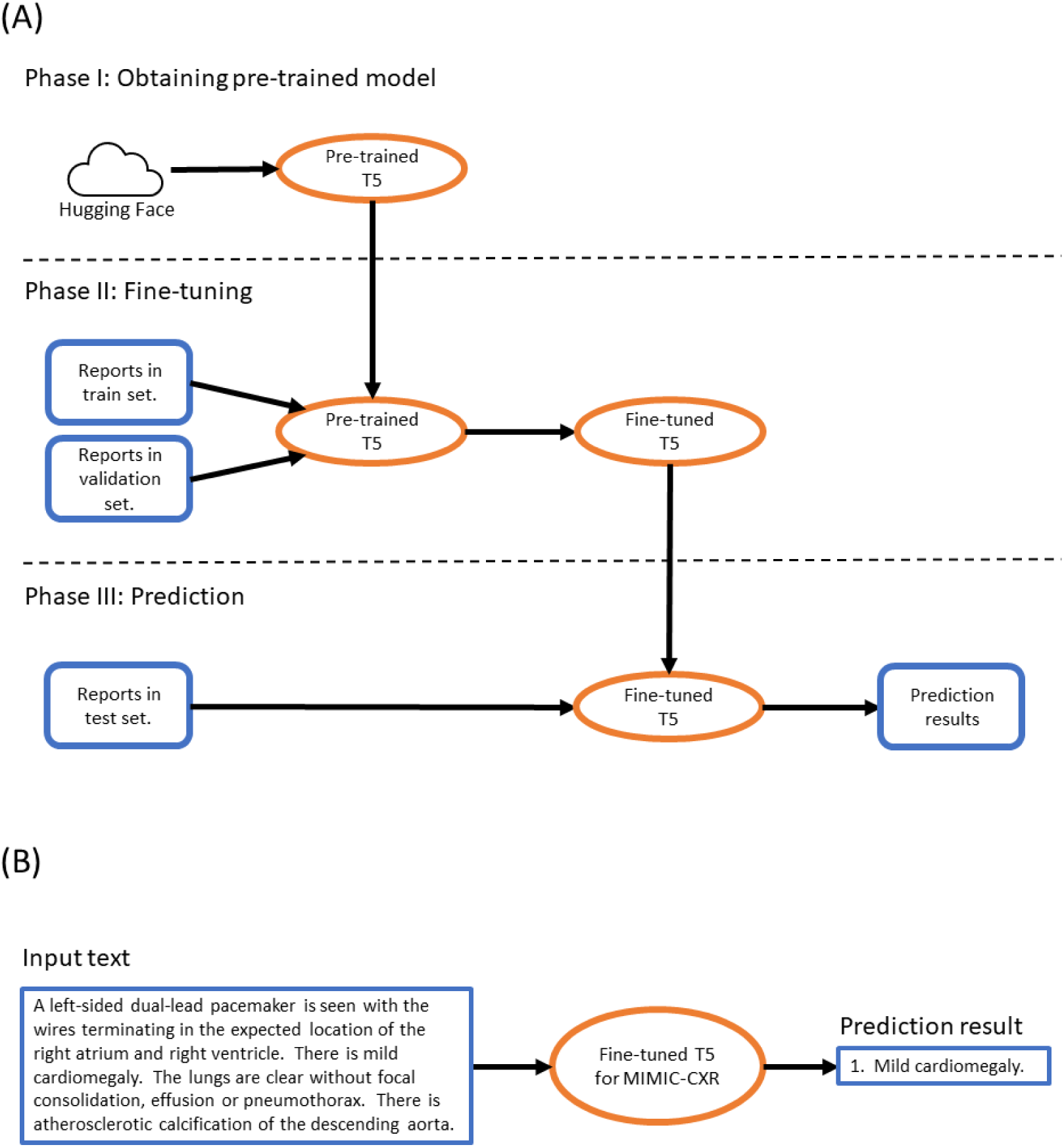
Outline of model development and prediction. (A) Flow of obtaining a pre-trained T5 model, fine-tuning the T5 model from the pre-trained model, and predicting the text of the impression section with the fine-tuned model. (B) Examples of the summary text predicted from the findings section. Abbreviations: T5, Text-to-Text Transfer Transformer

### Evaluation

The summarized text obtained from the fine-tuned T5 models was evaluated quantitatively using ROUGE metrics (21,22) and semi-quantitatively by radiologists. First, the ROUGE metrics were calculated between the impression text of the actual report and the predicted text. The ROUGE metrics indicate the summarization quality by measuring the alignment between the model-generated and original radiologist-generated summaries. The ROUGE metrics are detailed in the Appendix 2. Next, 100 reports were randomly selected from the test sets to undergo a semi-quantitative evaluation. Two radiologists with 17 and 7 years of experience in clinical radiology independently rated the predicted impression of the 100 reports on a 5-point scale as the semi-quantitative evaluation. The following text was evaluated: (i) the pairs of actual and predicted impression sections and (ii) the finding sections of the actual reports. The 5-point scores were defined as follows: 1, the predicted impression could not be used clinically without rewriting; 2, most of the predicted impressions requires rewriting to be clinically useful; 3, approximately half of the predicted impression requires rewriting to be clinically useful; 4, the predicted impression is clinically usable with minor modifications; and 5, the predicted impression is clinically usable without modification. The summarized text with scores of 4 and 5 was considered clinically useful. A consensus was reached through discussion in case of disagreements between the two radiologists.

### Statistics

The differences among the ROUGE metrics of the four T5 models were compared using the Wilcoxon signed-rank test. Quadratic-weighted kappa values were calculated for the scores of the two radiologists. The kappa values were interpreted using the following criteria: 0.00–0.20, none to slight; 0.21–0.40, fair; 0.41–0.60, moderate; 0.61–0.80, substantial; and 0.81–1.00, almost perfect agreement. Spearman’s correlation coefficients between the ROUGE-2 values and radiologists’ consensus scores were determined for the 100 reports of MIMIC-CXR and JMID, and the coefficients were statistically evaluated. Statistical significance was set at a p-value of 0.05. Statistical analyses were performed using R (version 4.2.2), Python (version 3.8.8), and Scipy (version 1.10.1).

## Results

### Dataset

Figure 1 presents the flowchart of the study. Table 1 presents the characteristics of the two datasets used in this study. Among the 227,781 reports present in the MIMIC-CXR dataset, 128,032 reports had no missing data. The findings or impression sections were frequently missing in the reports of the MIMIC-CXR dataset. The MIMIC-CXR dataset was divided into three subsets: a training set comprising 102,425 reports; a validation set comprising 6,402 reports; and a test set comprising 19,205 reports. The modality was radiography, and the location was specified as the chest in the MIMIC-CXR dataset, and the reports were written in English. Among the 1,120,311 reports in the JMID dataset, 1,101,271 had no missing data. The training, validation, and test sets comprised 1,035,574, 7,654, and 58,043 reports, respectively. The modalities in the JMID dataset were CT and MRI, with locations spanning different parts of the body, and the reports were written in Japanese.

**Table 1:** Characteristics of datasets.

| Item | MIMIC-CXR | JMID |
| --- | --- | --- |
| Number of reports | 227781 | 1120311 |
| Number of non-blank reports | 128032 | 1101271 |
| Number of reports in train set | 102425 | 1035574 |
| Number of reports in validation set | 6402 | 7654 |
| Number of reports in test set | 19205 | 58043 |
| Age (year) |  |  |
| train set | Not available | 62.33 ± 18.49 |
| validation set | Not available | 62.78 ± 18.16 |
| test set | Not available | 62.46 ± 18.82 |
| Sex (male:female) |  |  |
| train set | Not available | 555975:479599 |
| validation set | Not available | 4002:3652 |
| test set | Not available | 30463:27580 |
| Modality | X-ray | CT, MRI |
| Location | Chest | Various locations |
| Language | English | Japanese |
| Private/Public datasets | Public dataset | Private dataset |

Note: Non-blank reports indicate that neither the findings nor impression sections are blank in the radiology reports. Age was not available for 16754, 726, and 6210 reports in the training, validation, and test sets, respectively.
Abbreviations: JMID, Japan Medical Image Database

### Model performance

Table 2 presents the ROUGE metrics (ROUGE-1, ROUGE-2, and ROUGE-L) results of the fine-tuned T5 models in the MIMIC-CXR and JMID test sets. The results of the fine-tuned T5 models in the validation sets of MIMIC-CXR and JMID datasets are presented in the Appendix 3. Table 2 presents the ROUGE results for the four combinations of fine-tuned T5 models in the MIMIC-CXR and JMID datasets. The fine-tuned “t5-base” model with a batch size of 2 demonstrated the highest ROUGE-1, ROUGE-2, and ROUGE-L values (ROUGE-1 = 57.75 ± 30.99, ROUGE-2 = 49.96 ± 35.36, and ROUGE-L = 54.07 ± 32.48) in the MIMIC-CXR dataset. In contrast, the fine-tuned “google/mt5-base” model with a batch size of 8 achieved the lowest values (ROUGE-1 = 52.35 ± 31.36, ROUGE-2 = 43.97 ± 35.48, and ROUGE-L = 49.05 ± 32.53). The fine-tuned “google/mt5-base” model with a batch size of 2 demonstrated the highest ROUGE-1, ROUGE-2, and ROUGE-L values (ROUGE-1 = 50.00 ± 29.24, ROUGE-2 = 39.66 ± 30.21, and ROUGE-L = 47.87 ± 29.44) in the JMID dataset. In contrast, the fine-tuned “megagonlabs/t5-base-japanese-web” model with a batch size of 8 achieved the lowest values (ROUGE-1 = 44.30 ± 28.56, ROUGE-2 = 34.01 ± 28.44, and ROUGE-L = 42.11 ± 28.54). These results underscore the interaction between the type of pre-trained model, batch size, and dataset characteristics.

**Table 2:** ROUGE values in the test sets of MIMIC-CXR and JMID.

| Dataset | T5 model | ROUGE-1 | ROUGE-2 | ROUGE-L |
| --- | --- | --- | --- | --- |
| MIMIC-CXR | t5-base, B=2 | 57.75 ± 30.99 | 49.96 ± 35.36 | 54.07 ± 32.48 |
| MIMIC-CXR | t5-base, B=8 | 55.27 ± 30.55 | 46.98 ± 34.79 | 51.34 ± 31.99 |
| MIMIC-CXR | google/mt5-base, B=2 | 55.84 ± 31.26 | 47.86 ± 35.61 | 52.54 ± 32.59 |
| MIMIC-CXR | google/mt5-base, B=8 | 52.35 ± 31.36 | 43.97 ± 35.48 | 49.05 ± 32.53 |
| JMID | megagonlabs/t5-base-japanese-web, B=2 | 48.99 ± 29.49 | 38.80 ± 30.20 | 46.91 ± 29.64 |
| JMID | megagonlabs/t5-base-japanese-web, B=8 | 44.30 ± 28.56 | 34.01 ± 28.44 | 42.11 ± 28.54 |
| JMID | google/mt5-base, B=2 | 50.00 ± 29.24 | 39.66 ± 30.21 | 47.87 ± 29.44 |
| JMID | google/mt5-base, B=8 | 46.18 ± 28.39 | 35.69 ± 28.65 | 43.91 ± 28.46 |

Note: Values represent the mean ± standard deviation. The numbers of reports are 19205 and 58043 for the test sets of MIMIC-CXR and JMID, respectively. Abbreviations: T5, Text-to-Text Transfer Transformer; B, batch size; ROUGE, Recall-Oriented Understudy for Gisting Evaluation.

The differences in the ROUGE-1, ROUGE-2, and ROUGE-L values were statistically evaluated between each pair of the four fine-tuned models. The p-values of ROUGE-1, ROUGE-2, and ROUGE-L were <.001 in the pairs of the four models in the MIMIC-CXR dataset, except for the p-values of ROUGE-1 between “google/mt5-base” with a batch of 2 and “t5-base” with a batch of 8. The p-values of ROUGE-1 between “google/mt5-base” with a batch of 2 and “t5-base” with a batch of 8 was 0.12. As the number of test set was larger in JMID dataset than that in MIMIC-CXR dataset, the p-values of ROUGE-1, ROUGE-2, and ROUGE-L were <.001 for each pair of the four models in JMID. Thus, we focused on the optimal fine-tuned models (the fine-tuned “t5-base” model with a batch size of 2 for MIMIC-CXR and the fine-tuned “google/mt5-base” model with a batch size of 2 for JMID) based on the p-values.

Table 3 presents the results of the radiologists’ semi-quantitative scores for the predicted summaries of the 100 reports generated by the optimal fine-tuned models of MIMIC-CXR and JMID. The kappa values of the semi-quantitative scores between the two radiologists were 0.785 (95% confidence interval = 0.669–0.900) and 0.736 (95% confidence interval = 0.590–0.883) for the 100 reports acquired from the MIMIC-CXR and JMID test sets, respectively, indicating substantial agreement between the two radiologists. The number of reports for the consensus scores was as follows: score 1 = 1, score 2 = 2, score3 = 11, score 4 = 15, and score 5 = 71 in MIMIC-CXR; score 1 = 2, score 2 = 3, score3 = 10, score 4 = 25, and score 5 = 60 in JMID. These results indicate that 86% (86/100) and 85% (85/100) of the automatically summarized texts were clinically useful in the MIMIC-CXR and JMID datasets, respectively.

**Table 3:** Results of semi-quantitative evaluation by radiologists in 100 radiology reports of test sets.

| Item | MIMIC-CXR | JMID |
| --- | --- | --- |
| Number of reports evaluated by radiologists | 100 | 100 |
| Number of Score 1 | 1 | 2 |
| Number of Score 2 | 2 | 3 |
| Number of Score 3 | 11 | 10 |
| Number of Score 4 | 15 | 25 |
| Number of Score 5 | 71 | 60 |

Note: Scores were determined by the consensus of the two radiologists. Abbreviations: JMID, Japan Medical Image Database

Figure 3 presents the scatter plots illustrating the relationships between the ROUGE-2 values and semi-quantitative scores of the two radiologists. Significant positive correlations were observed between the ROUGE-2 values and semi-quantitative scores in both datasets. The calculated correlation coefficients were 0.446 (95% confidence interval = 0.274–0.591) and 0.261 (95% confidence interval = 0.0681–0.435) for the MIMIC-CXR and JMID datasets, respectively. The corresponding p-values of the correlation coefficients were <.001 for both datasets, indicating that the positive correlations observed between the ROUGE-2 values and the semi-quantitative scores were statistically significant. Scatter plots of the ROUGE-2 values and semi-quantitative scores for each of the two radiologists are presented in the Appendix 4 and 5. Figure 4 presents representative examples of radiology reports and the summary text predicted by the fine-tuned T5 models.

**Figure 3.**
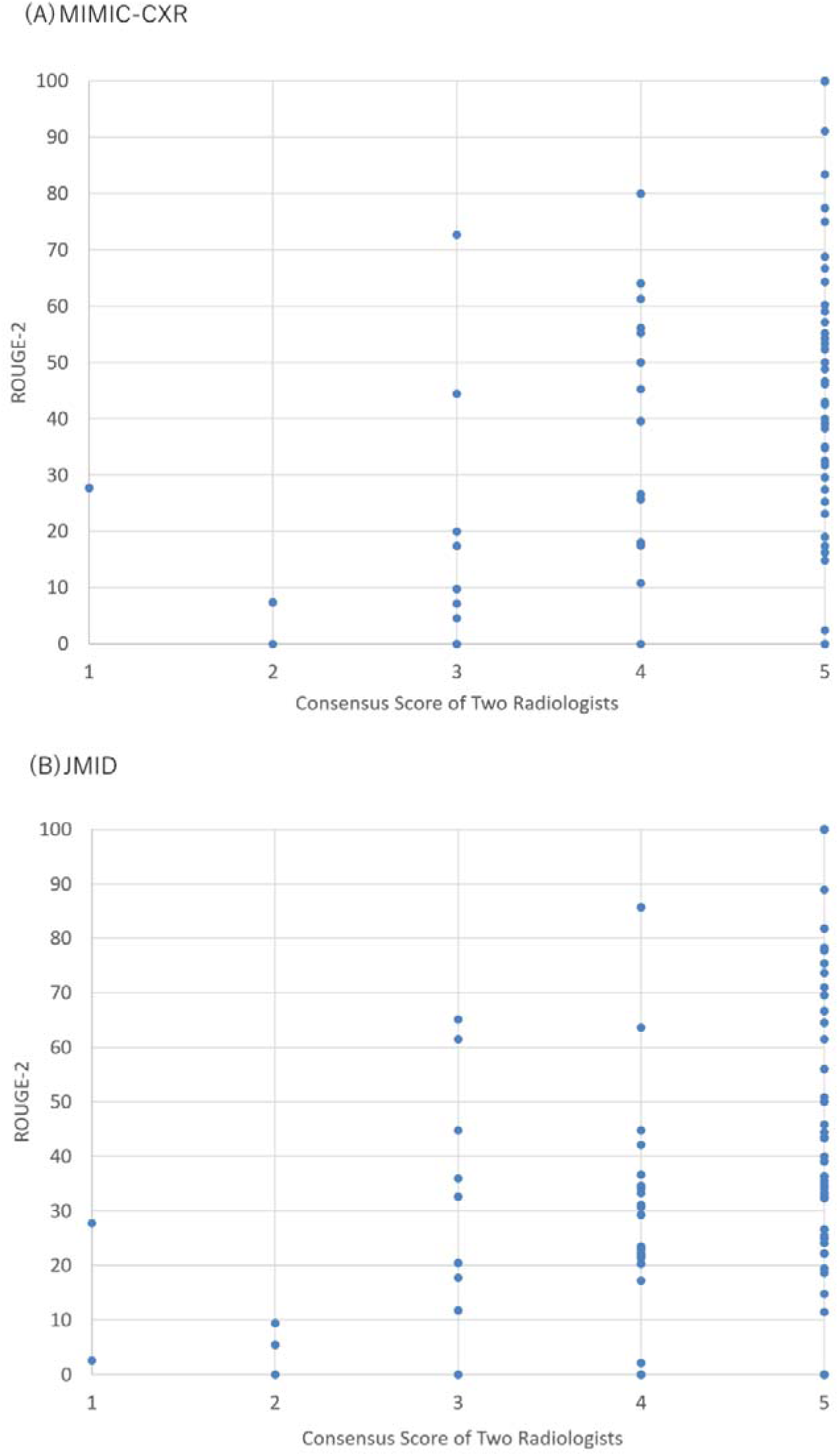
Scatter plots between the ROUGE-2 values and semi-quantitative consensus scores by two radiologists. (A) Scatter plot for MIMIC-CXR, (B) Scatter plot for JMID. Note for (A): Correlation coefficient and p-value between ROUGE-2 values and consensus scores were 0.446 and <.001, respectively. Note for (B): Correlation coefficient and p-value between ROUGE-2 values and consensus scores were 0.261 and <.001, respectively. Abbreviations: JMID, Japan Medical Image Database; ROGUE, Recall-Oriented Understudy for Gisting Evaluation

**Figure 4.**
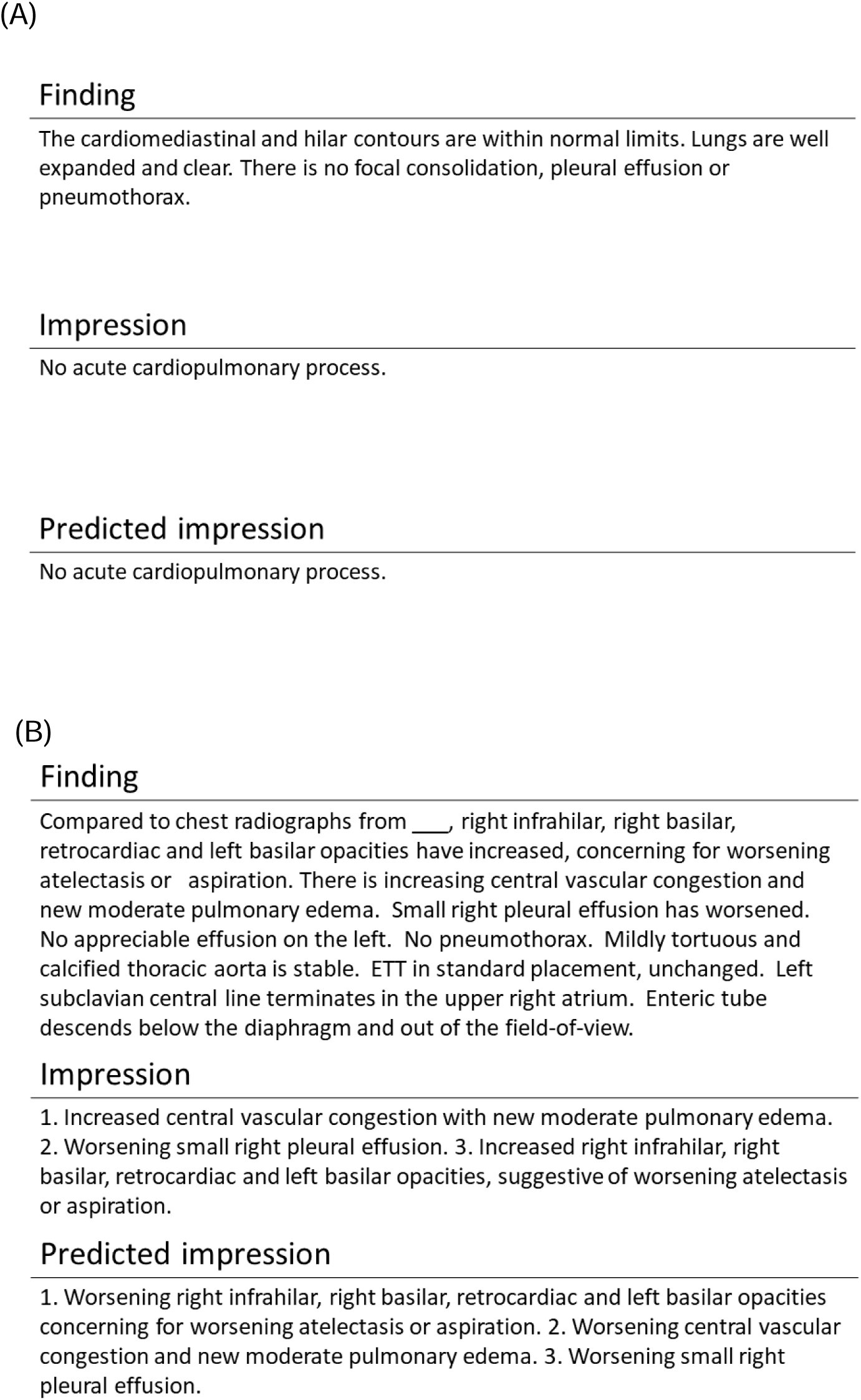
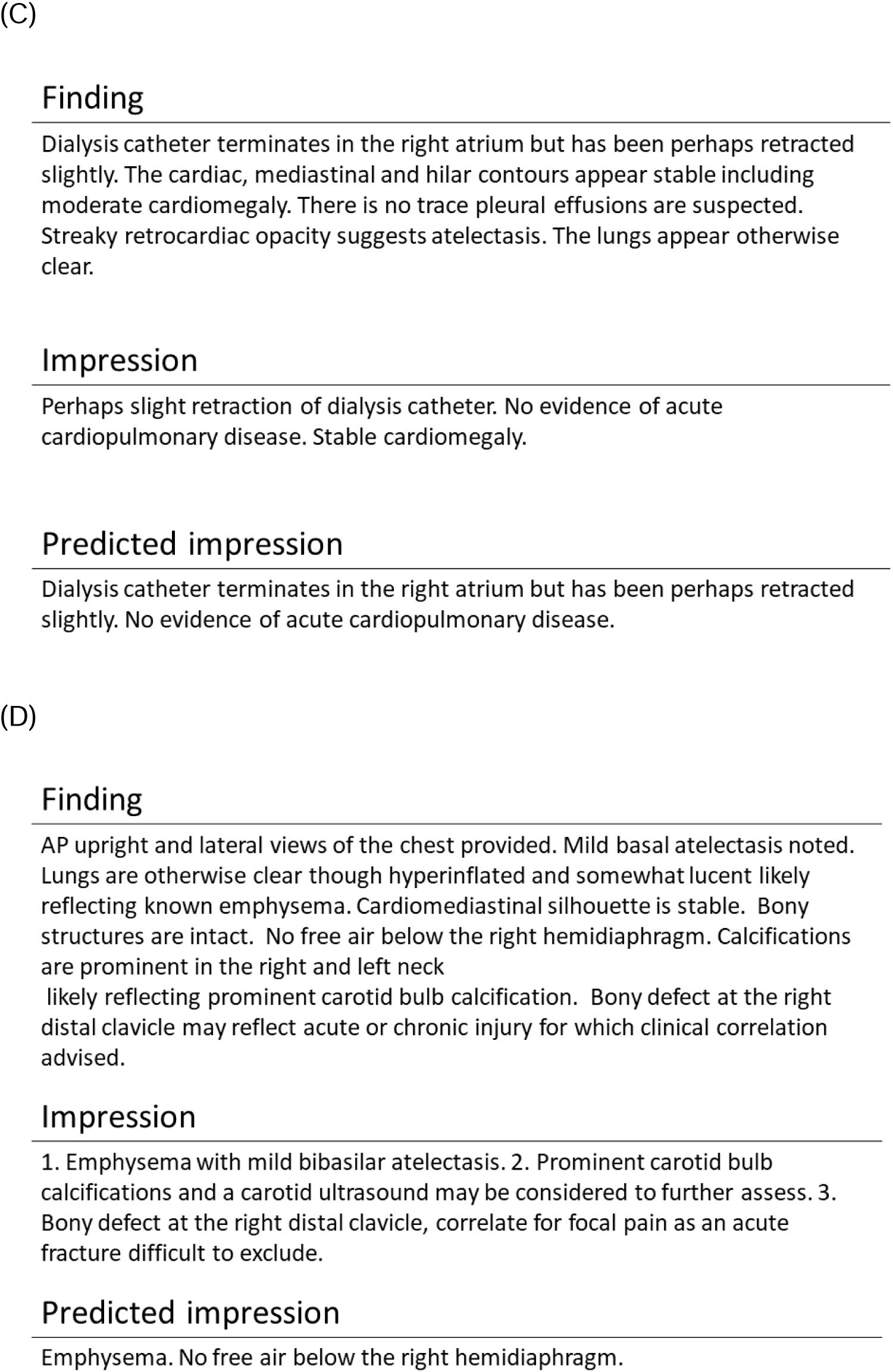
Representative example of radiology reports and predicted summary text from MIMIC-CXR. Note: The radiologists’ scores are as follows: A = 5, B = 5, C = 4, D = 2.

## Discussion

An automatic summarization model of NLP was constructed using the T5 model. The fine-tuned T5 model was capable of summarizing radiology reports automatically. The present study revealed that the fine-tuned “t5-base” model with a batch size of 2 for MIMIC-CXR and the fine-tuned “google/mt5-base” model with a batch size of 2 for the JMID were the best T5 models. The scores of the radiologists’ semi-quantitative evaluation of the 100 reports were ≥4 for 86% in the MIMIC-CXR test set and 85% in the JMID test set. This indicates that most of the predicted text in the impression section was clinically useful. These results demonstrate the usefulness of the T5 summarization models in the automated summarization of radiology reports. In addition, statistically significant correlations were observed between the radiologists’ semi-quantitative scores and the quantitative evaluation using ROUGE metrics for the MIMIC-CXR and JMID datasets.

MIMIC-CXR is a dataset comprising reports and CXR images (15); however, the anatomical locations and diseases included in MIMIC-CXR are relatively limited. In contrast, the JMID dataset comprises CT and MR images of all locations, and the anatomical locations and diseases involved are broader than those in MIMIC-CXR. Consequently, report variation was greater in the JMID dataset, and report summarization was more difficult in JMID than in MIMIC-CXR. Table 3 demonstrates that the radiologists’ scores were comparable for the two datasets. JMID comprised a greater number of reports, approximately ten times more than MIMIC-CXR (Table 1). The results presented in Tables 1 and 3 suggest that the number of radiology reports influenced the performance of the automatic summarization models. Thus, the dataset size for constructing language models may be important in NLP, similar to computer vision (23,24).

The two different pre-trained T5 models were used for MIMIC-CXR and JMID in the present study: “t5-base” and “google/mt5-base” for MIMC-CXR; “megagonlabs/t5-base-japanese-web” and “google/mt5-base” for JMID. The best ROUGE values were obtained with “t5-base” for MIMIC-CXR and “google/mt5” for JMID (Table 2). This result indicates that the English T5 model outperformed the multilingual T5 model in the English task, whereas the multilingual T5 model outperformed the Japanese T5 model in the Japanese task. In general, considering the variation in dataset sizes based on the languages, English stands out owing to its larger size. The results of the present study and the dataset size of English datasets suggest that utilizing models pre-trained in English datasets would be more effective for English tasks because the pre-trained model specialized for English has sufficient generalizability. However, the dataset size used for pre-training was smaller for languages other than English. Consequently, the multilingual model would be more effective for non-English tasks because the multilingual model was pre-trained using both English and non-English datasets (8).

Similar to a previous study (21), statistically significant correlations were observed between the ROUGE-2 values and the radiologists’ semi-quantitative scores for the 100 reports selected from the test sets. The Spearman’s correlation coefficients were 0.446 for MIMIC-CXR and 0.261 for JMID, indicating weak correlations. The ROUGE-2 values exhibited significant variability even when the radiologists’ score was 5 (Figure 3). Therefore, evaluating individual summarized sentences by relying solely on the ROUGE metrics may not be reliable. However, the average ROUGE values could potentially serve as a surrogate for the average radiologist scores when the test set (evaluation dataset) is sufficiently large. These two evaluation methods should be used complementarily, particularly when the evaluation dataset is limited.

JMID is a large dataset with a size of more than one million. Creating a dataset larger than that of the JMID is difficult. Conversely, to achieve significant improvements in the performance of automatic summarization, significant improvements are required in the summarization model architecture or the pre-trained model.

This study had certain limitations. First, we speculate that the usefulness of a multilingual or language-specific pre-trained model depends on the language of the radiology reports. However, this may depend on the dataset characteristics other than the language. As only two datasets were utilized, this aspect could not be adequately evaluated in this study. Second, only two languages were used in this study. Other languages should be investigated in future studies. Third, the dataset sizes of MIMIC-CXR and JMID were relatively large for medical NLP. Smaller datasets were not used in this study.

In conclusion, this study demonstrated the feasibility of automatic report summarization, and the majority of the automatically summarized text was clinically useful. Significant correlations were observed between the semi-quantitative evaluations performed by radiologists and the quantitative evaluations by the ROUGE metrics. The use of quantitative assessments provided by the ROUGE metrics in conjunction with the semi-quantitative scores provided by radiologists could be complementary. These results could lead to further advances in NLP radiology research.

## Data Availability

All data produced in the present study are available upon reasonable request to the corresponding author.

## Acknowledgement

We thank JMID project and the following 10 academic medical centers.

- Juntendo University
- Kyushu University
- Keio University
- The University of Tokyo
- Okayama University
- Kyoto University
- Osaka University
- Hokkaido University
- Ehime University
- Tokushima University

## Abbreviations

CXR: Chest x-ray
JMID: Japan Medical Image Database
NLP: Natural language processing
ROUGE: Recall-Oriented Understudy for Gisting Evaluation
T5: Text-to-Text Transfer Transformer

## Appendix 1 Details of Text-to-Text Transfer Transformer (T5) model

Text-to-Text Transfer Transformer (T5) is a transformer-based deep learning model that uses a text-to-text approach. Most NLP tasks, including translation, question answering, and classification, comprise giving input sentences to models and training them to generate target sentences. This facilitates the use of the same model, loss function, and hyperparameters for a variety of tasks. The T5 has model achieved state-of-the-art results on many NLP benchmarks, while maintain sufficient flexibility to be fine-tuned for a variety of important downstream tasks.

A large amount of unlabeled text and an objective analogous to BERT’s “masked language modeling” were used to pre-train the T5 model. Tokens of input text were randomly corrupted with special tokens during pre-training. The pre-trained T5 reconstructed the corrupted tokens of the input text after pre-training.

the pre-trained T5 model can be used for a variety of text-to-text tasks; however, text summarization of radiology reports was performed as a downstream task in this study. The following pre-trained T5 models were used to summarize the reports in the MIMIC-CXR and JMID datasets.

- MIMIC-CXR: t5-base or google/mt5-base
- JMID: megagonlabs/t5-base-japanese-web or google/mt5-base

The t5-base, megagonlabs/t5-base-japanese-web, and google/mt5-base were pre-trained with unlabeled English, Japanese, and multilingual text, respectively.

A character encoding conversion to UTF-8 was performed for the Japanese text of JMID as a preprocessing step before fine-tuning the T5 models. No preprocessing was performed for MIMIC-CXR.

The following hyperparameters were used to fine-tune the pre-trained T5 models.

- Number of tokens in input: 1024
- Number of tokens in output: 128
- Number of training epochs: 5
- Batch size: 2 or 8
- Number of steps for gradient accumulation: 32
- Learning rate: 5e-5
- Learning scheduler: cosine annealing
- Warmup ratio in cosine annealing: 0.05

The following parameter was used to generate the predicted impression using the fine-tuned T5 models.

- Number of tokens in beam search: 6

Python (version, 3.8.8), pytorch (version, 1.8.0), and transformers (version, 4.22.2) were used for model development. The T5 models were developed and evaluated on a workstation with NVIDIA(R) RTX(TM) A6000.

For the model development and summary prediction, run_summarization.py of transformers was used in the present study (https://github.com/huggingface/transformers/blob/main/examples/pytorch/summarization/run_summarization.py).

## Appendix 2 Details of ROUGE

ROUGE is a commonly used metric that is used to evaluation of text summarization. This metric measures the alignment between human-generated summaries (reference summaries) and summaries generated by a model. ROUGE has several variants. ROUGE-1, ROUGE-2, and ROUGE-L were used in this study. ROUGE-1 and ROUGE-2 are basic metrics that measure the alignment on an n-gram basis. ROUGE-1 and ROUGE-2 use unigrams and bigrams, respectively. The original definition of ROUGE-1 and ROUGE-2 is as follows:

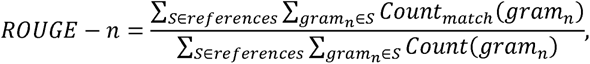

where *n* represents the length of the n-gram and Count_match_ (gram_n_) represents the maximum number of n-gram co-occurring in a model-generated summary and a reference summary. Three options can be used to calculate ROUGE-1 and ROUGE-2: recall basis, precision basis, and F-measure basis. F-measure-based ROUGE-1 and ROUGE-2 were used in this study.

ROUGE-1 and ROUGE-2 use the frequency of n-gram co-occurring; however, ROUGE-L uses common subsequence between human-generated summaries and model-generated summaries. Given two sequences *X* and *Y* (where *X* is a reference summary sentence, and *Y* is a model-generated summary sentence), the longest common subsequence (LCS) of *X* and *Y* is defined as a common subsequence with maximum length. To estimate the similarity between two summaries *X* of length *m* and *Y* of length *n*, ROUGE-L (F_lcs_) is defined as LCS-based F-measure according to the following equations:

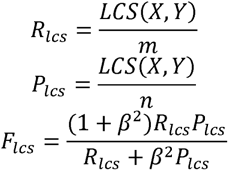

Summaries must be tokenized for the evaluation of the alignment in calculating ROUGE metrics. BERT’s tokernizer was used to perform tokenization in this study.

## Appendix 3

Validation loss of MIMIC-CXR and JMID in the fine-tuning of the T5 models.

| Dataset | T5 model | Validation loss |
| --- | --- | --- |
| MIMIC-CXR | t5-base, B=2 | 1.0537 |
| MIMIC-CXR | t5-base, B=8 | 1.1476 |
| MIMIC-CXR | google/mt5-base, B=2 | 1.0431 |
| MIMIC-CXR | google/mt5-base, B=8 | 1.1693 |
| JMID | megagonlabs/t5-base-japanese-web, B=2 | 0.9893 |
| JMID | megagonlabs/t5-base-japanese-web, B=8 | 1.098 |
| JMID | google/mt5-base, B=2 | 0.9422 |
| JMID | google/mt5-base, B=8 | 1.0344 |

Note: Validation sets of MIMIC-CXR and JMID comprised 6402 and 7654 reports, respectively.
Abbreviations: T5, Text-to-Text Transfer Transformer; B, batch size.

## Appendix 4

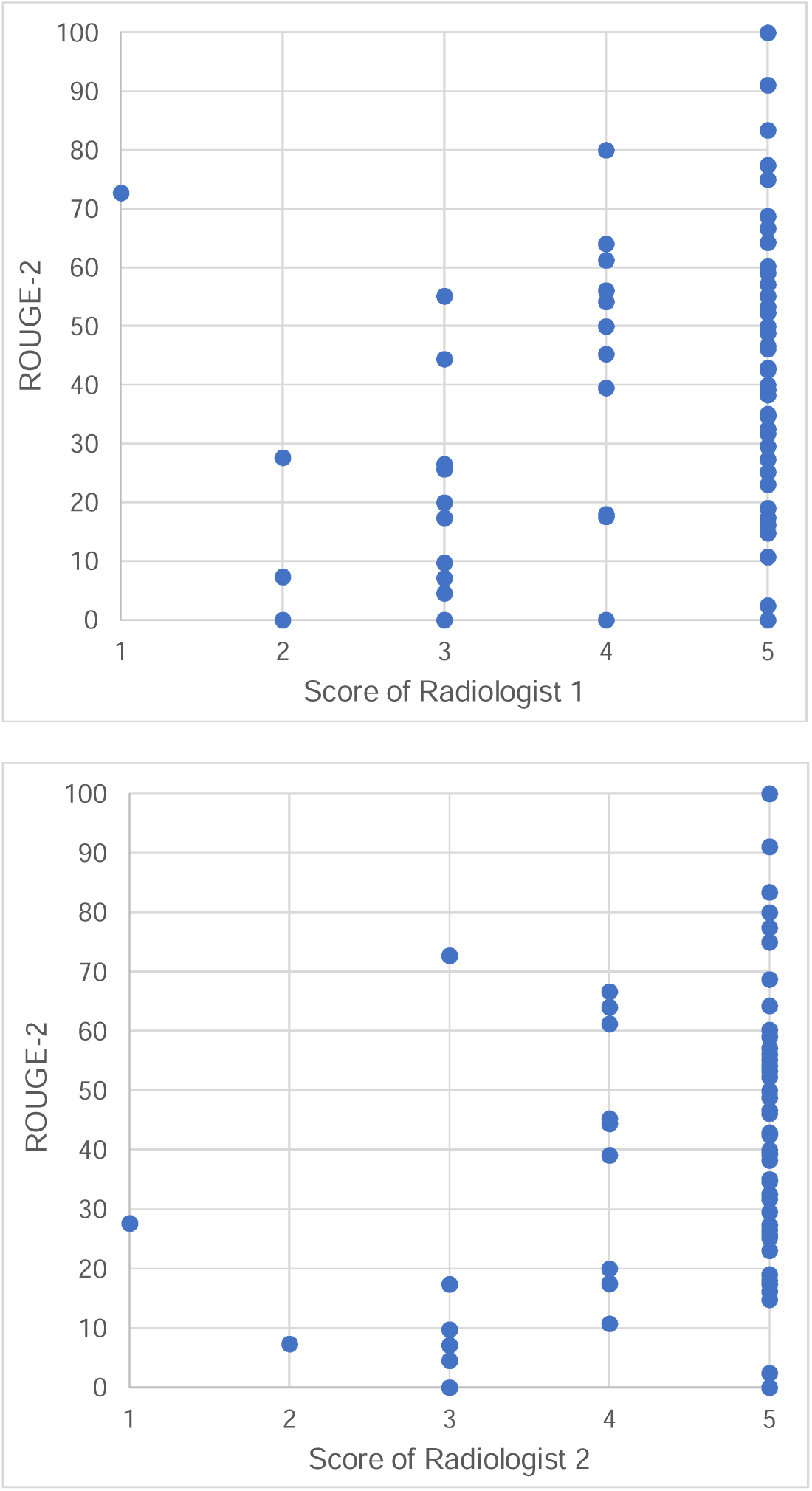

Scatter plots of the ROUGE-2 values and radiologist’ semi-quantitative scores in MIMIC-CXR.

## Appendix 5

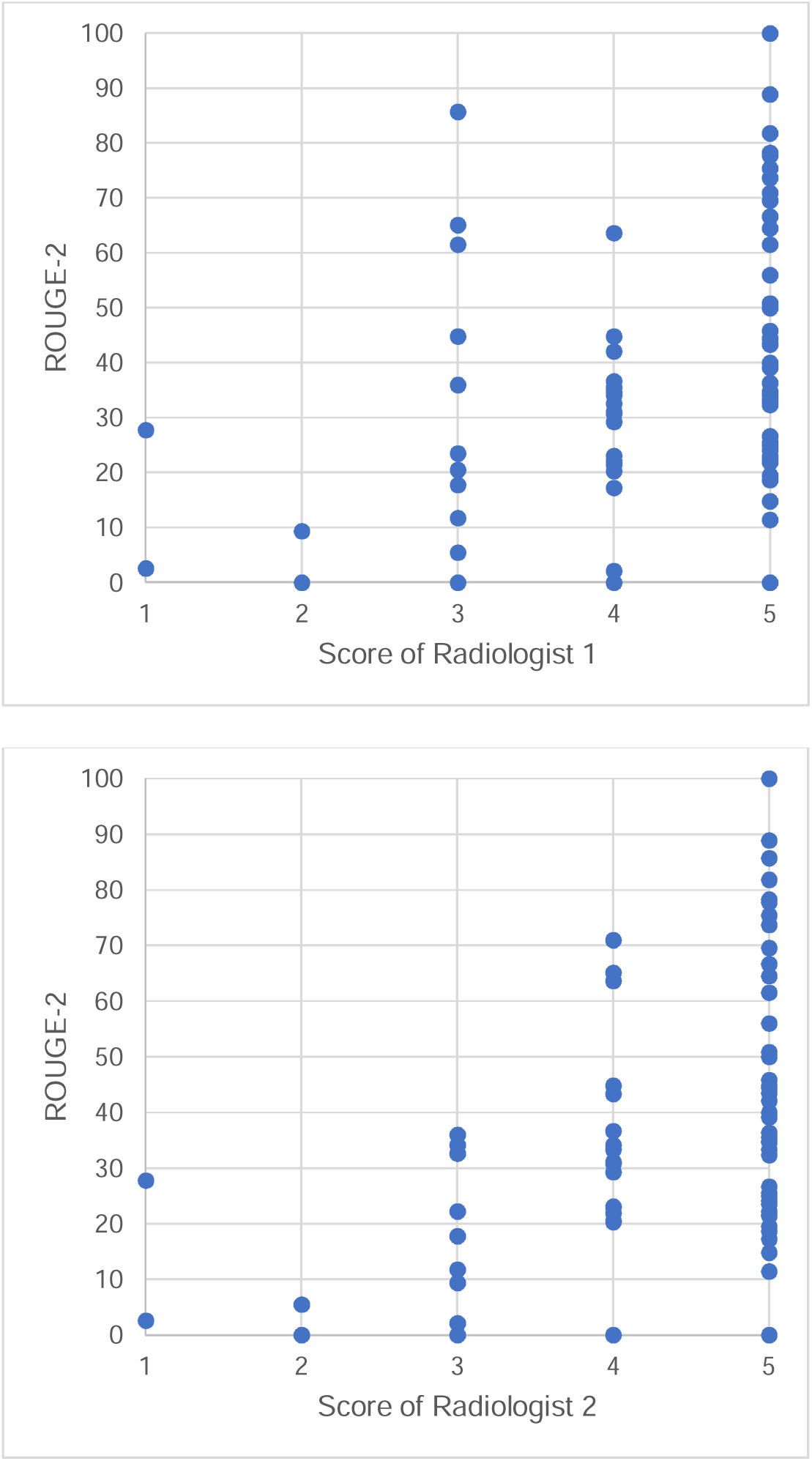

Scatter plots of the ROUGE-2 values and radiologist’ semi-quantitative scores in JMID.

